# Employing Consensus-Based Reasoning with Locally Deployed LLMs for Enabling Structured Data Extraction from Surgical Pathology Reports

**DOI:** 10.1101/2025.04.22.25326217

**Authors:** Aakash Tripathi, Asim Waqas, Kavya Venkatesan, Ehsan Ullah, Asma Khan, Farah Khalil, Wei-Shen Chen, Zarifa Gahramanli Ozturk, Daryoush Saeed-Vafa, Marilyn M. Bui, Matthew B. Schabath, Ghulam Rasool

## Abstract

Surgical pathology reports provide essential diagnostic information critical for cancer staging, treatment planning, and cancer registry documentation. However, their writing styles and formats vary widely, reflecting each pathologist’s stylistic choices, institutional norms, and largely inherited practices from residency training. When performing large-scale data analysis, this unstructured nature and variability across tumor types and institutions pose significant hurdles for automated data extraction. To overcome these challenges, we present a consensus-driven, reasoning-based framework that adapts multiple locally deployed large language models (LLMs) to extract both standard diagnostic variables (site, laterality, histology, stage, grade, and behavior) and organ-specific biomarkers. Each LLM generates structured outputs, accompanied by justifications, which are subsequently evaluated for accuracy and coherence by three separate reasoning models (DeepSeek-R1-large, Qwen3-32B, and QWQ-32B). Final consensus values are determined through aggregation. Board-certified pathologists conducted expert validation. This framework was applied to over 6,100 pathology reports from The Cancer Genome Atlas (TCGA) spanning 10 organ systems and 510 reports from Moffitt Cancer Center. For the TCGA dataset, automated evaluation demonstrated mean accuracy of 84.9% ± 7.3%, with histology (89.0%), site (88.3%), and behavior (87%) showing the highest extraction accuracy averaged across all models. Expert review of randomly selected 138 reports confirmed high agreement for behavior (100.0%), histology (99%), grade (97%), and site (95%) in the TCGA dataset, with slightly lower performance for stage (88%) and laterality (87%). In Moffitt Cancer Center reports (brain, breast, and lung), accuracy remained high (88.2% ± 7.2%), with behavior (99%), histology and laterality (96%), grade (93%), and site (91%) achieving strong agreement. Biomarker extraction achieved 70.6% ± 8.1% overall accuracy, with TP53 (85%) on brain tumor, Ki-67 (68%) on breast cancer, and ROS1 (82%) on lung cancer showing highest accuracy. Inter-evaluator agreement analysis revealed high concordance (correlations > 0.89) across the three evaluation models. Statistical analyses revealed significant main effects of model type (F=1716.82, p<0.001), variable (F=3236.68, p<0.001), and organ system (F=1946.43, p<0.001), as well as model × variable × organ interactions (F=24.74, p<0.001), emphasizing the role of clinical context in model performance. These results highlight the potential of stratified, multi-organ evaluation frameworks with multi-evaluator consensus in LLM benchmarking for clinical applications. Overall, this consensus-based approach demonstrates that locally deployed LLMs can provide a transparent, accurate, and auditable solution for integration into real-world pathology workflows such as synoptic reporting and cancer registry abstraction.

## 1 Introduction

Surgical pathology reports are the cornerstone of cancer diagnosis, staging, and histologic classification. Additionally, they provide valuable information to guide treatment selection, inform prognostic assessments, and determine eligibility for clinical trials [1, 2, 3]. The information contained in these reports also underpin national cancer surveillance efforts, such as the Surveillance, Epidemiology, and End Results (SEER) program and the National Program for Cancer Registries (NPCR), which require consistent capture of variables like tumor site, laterality, histological diagnosis, stage, grade, and behavior [4, 5, 6, 7, 8]. Despite their centrality, most pathology reports are composed as unstructured free-text narratives, often dictated in formats that vary widely within and across institutions, subspecialties, and pathologists [9]. This heterogeneity poses significant challenges for automated information extraction and downstream reuse. While rule-based natural language processing (NLP) systems have been applied to map free-text to discrete fields, they are time-consuming to develop, sensitive to linguistic variation, and poorly generalize across diverse cancer types and documentation styles [9, 10, 11, 12]. As the holistic cancer care and research increasingly depend on large-scale, structured datasets to power precision medicine and population health efforts, there is a critical and urgent need for extraction methods that are not only accurate and scalable but also adaptable to real-world documentation variability.

Recent advances in large language models (LLMs), including ChatGPT, LLaMA, and DeepSeek, have enabled automated extraction of structured information from free-text clinical documents, including radiology, discharge summaries, and pathology reports [13, 14, 15, 16, 17, 18, 8, 19, 20]. These models are capable of identifying key clinical entities and relationships with little or no task-specific training, making them candidates for applications that require scalability across cancer types and report formats. However, despite their linguistic fluency, LLMs suffer from key limitations that restrict their reliability in clinical workflows. A major concern is their tendency to produce plausible-sounding but factually incorrect or unsupported outputs, a phenomenon often referred to as hallucination. [21, 22, 23]. Hallucinations may result in fabricated stage values, incorrect site mappings, or invalid combinations of diagnostic attributes, all of which pose risks in settings like cancer registry abstraction or clinical trial matching. In addition, most LLMs do not express uncertainty and may provide definitive-sounding answers even when source evidence is weak or contradictory. These behaviors, combined with the opacity of current LLM decision-making processes, have raised concerns among clinicians and pathologists about their safety and trustworthiness [24, 25, 26, 27]. As a result, there is a growing consensus that LLM-based extraction systems must incorporate structured reasoning, interpretability, and safeguards that mitigate error propagation in high-stakes clinical contexts.

Despite their promise, the deployment of LLMs in clinical environments is constrained by stringent requirements around patient privacy, regulatory compliance, and institutional data governance. Cloud-based LLMs, such as those offered by commercial providers, often require transmitting protected health information to external servers, raising concerns about data breaches and non-compliance with regulations like HIPAA and GDPR [28]. These concerns are heightened by recent controversies around unauthorized model training on clinical data, leading to increased institutional scrutiny and legal risk. As a result, there is growing interest in deploying LLMs locally within secure hospital infrastructures, where data remains behind institutional firewalls and access can be tightly controlled [29, 30, 31]. Locally hosted models also offer greater transparency, auditability, and customization for clinical use cases, particularly when based on open-source architectures. However, practical challenges remain: most LLMs require substantial computational resources, and integration into clinical workflows demands interoperability with legacy systems, domain-specific prompt design, and support for human oversight. Addressing these barriers is critical to translate LLMs from research prototypes to deployable clinical tools.

To bridge the gap between the capabilities of LLMs and the demands of clinical pathology, we developed a consensus-based framework that emphasizes accuracy, transparency, and clinical alignment. Rather than relying on a single model to interpret free-text reports, our approach leverages multiple locally deployed LLMs, each prompted to extract discrete diagnostic variables such as tumor site, histology, and behavior, alongside concise textual justifications for their selections. A separate reasoning model then evaluates these outputs, synthesizes inter-model agreement, and adjudicates conflicts based on justification strength and internal consistency. This multi-stage process mirrors how diagnostic consensus is achieved in clinical practice: through independent interpretation, reasoned comparison, and reconciliation of divergent opinions. By embedding structured reasoning into every stage of prediction and adjudication, the system produces results that are not only accurate but also explainable and auditable, which are the qualities essential for downstream integration into cancer registries, synoptic reporting systems, and clinical trial eligibility assessments.

This work distinguishes itself by combining locally deployed LLMs with a reasoning-based consensus architecture that prioritizes auditability, transparency, and clinical alignment. Rather than producing black-box outputs, the system generates structured predictions with accompanying justifications and resolves conflicts through an adjudication step based on coherence, agreement, and clinical plausibility. These features enable human review, support regulatory compliance, and foster trust in high-stakes environments like cancer registry abstraction and synoptic reporting. Designed to run entirely within institutional firewalls using quantized open-source models, the framework can be deployed on modest hardware, making it scalable and compatible with existing pathology workflows.

In this study, we apply a consensus-based reasoning framework to two large corpora of surgical pathology reports: a publicly available dataset from The Cancer Genome Atlas (TCGA) and a real-world institutional dataset from Moffitt Cancer Center, comprised of breast, brain, and lung cancers. The system operates entirely on locally deployed infrastructure, using multiple open-source LLMs to extract six clinically relevant variables along with model-generated justifications. A reasoning module adjudicates conflicting outputs, producing final consensus predictions that are auditable and suitable for human review. Model performance is evaluated using automated metrics and expert validation by board-certified or equivalent pathologists, and we conduct detailed statistical analyses to examine model accuracy across organ systems and diagnostic fields. Together, these findings demonstrate the clinical feasibility of deploying AI-driven extraction tools in routine pathology workflows while highlighting the importance of interpretability, adjudication, and dataset diversity in building trustworthy systems for structured data abstraction.

## 2 Methods

A locally deployable, three-stage framework was developed for extracting structured clinical variables from unstructured surgical pathology reports using LLMs, as shown in Figure 1. The system targets six key diagnostic variables, including site, laterality, histology, stage, grade, and behavior, and emphasizes interpretability by requiring each model to produce brief justifications alongside its predictions. The framework integrates outputs from multiple LLMs, evaluates their accuracy, and applies reasoning-based aggregation to produce a final consensus output [32]. To validate its reliability and clinical relevance, we applied the framework to two diverse pathology datasets and conducted both automated evaluation and expert pathologist review.

**Figure 1:**
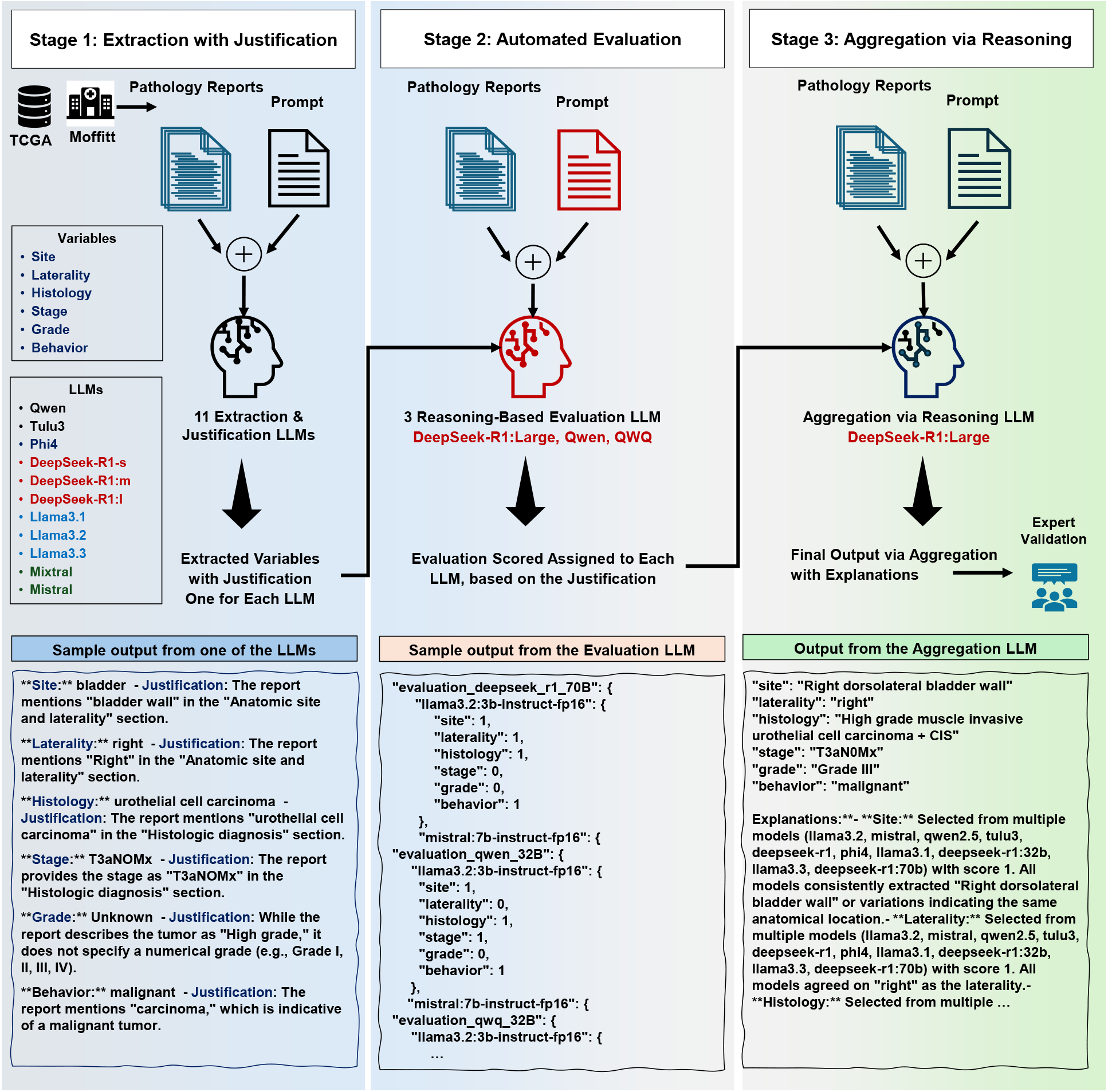
Multi-stage reasoning framework for structured information extraction from surgical pathology reports. This diagram illustrates the full three-stage prompting pipeline developed to extract six variables, site, laterality, histology, stage, grade, and behavior, from unstructured pathology reports. Reports were sourced from two datasets: TCGA and Moffitt Cancer Center, encompassing eight organ types across more than 4,000 reports. In **Stage 1**, eleven locally hosted LLMs (LLaMA 3.1, LLaMA 3.2, LLaMA 3.3, Mistral, Mixtral-medium, Qwen, Phi-4-medium, Tulu3-small, DeepSeek-R1:7B, DeepSeek-R1-medium, and DeepSeek-R1-large) were independently prompted to extract variable values and provide textual justifications. In **Stage 2**, DeepSeek-R1-large, QWQ, and Qwen were used as an automated evaluator to assign binary accuracy scores (1 = correct, 0 = incorrect) to each model’s predictions by comparing them to the original report and corresponding justification. In **Stage 3**, a second prompt to DeepSeek-R1-large served as an aggregation step, integrating all model predictions and their evaluation scores to select final consensus values. Outputs from this stage included not only the selected variable values but also concise rationales explaining the model’s reasoning. A subset of outputs from both datasets was randomly sampled and independently reviewed by board-certified or equivalent pathologists, who scored each extracted variable and annotated common error patterns. This framework enables interpretable, accurate, and auditable structured data extraction suitable for integration into real-world clinical workflows such as cancer registry abstraction and synoptic reporting.

### 2.1 Prompting and Reasoning Framework

In the first stage, each pathology report was independently processed by eleven locally deployed LLMs (detailed specifications provided in Table 1): LLaMA 3.1:8B (LLaMA 3.1-small), LLaMA 3.2:3B (LLaMA 3.2-small), LLaMA 3.3:70B (LLaMA 3.3-large), Mistral-7B (Mistral-small), Mixtral-8x7B (Mixtral-medium), Qwen 2.5:7B (Qwen-small), Phi-4:14B (Phi-medium), Tulu3:8B (Tulu-small), DeepSeek-R1:8B (DeepSeek-R1-small), DeepSeek-R1:32B (DeepSeek-R1-medium), and DeepSeek-R1:70B (DeepSeek-R1-large). Each model received the full report text and was prompted to extract six structured clinical variables along with a brief justification for each value. For the Moffitt biomarker cohort, models additionally extracted organ-specific biomarkers: breast (ER, PR, HER2, Ki-67), brain (IDH1/2, MGMT, ATRX, TP53, 1p/19q codeletion, TERT, EGFR, CDKN2A/B), and lung (PD-L1, EGFR, ALK, ROS1, BRAF, KRAS, MET, RET, HER2, NTRK). If a variable was absent or could not be inferred, the model was instructed to return “Unknown” with an explanation. Outputs were returned in standardized JSON format to support downstream scoring and aggregation. For brain cancer reports, the “stage” variable was excluded due to the lack of clinical relevance.

**Table 1:**
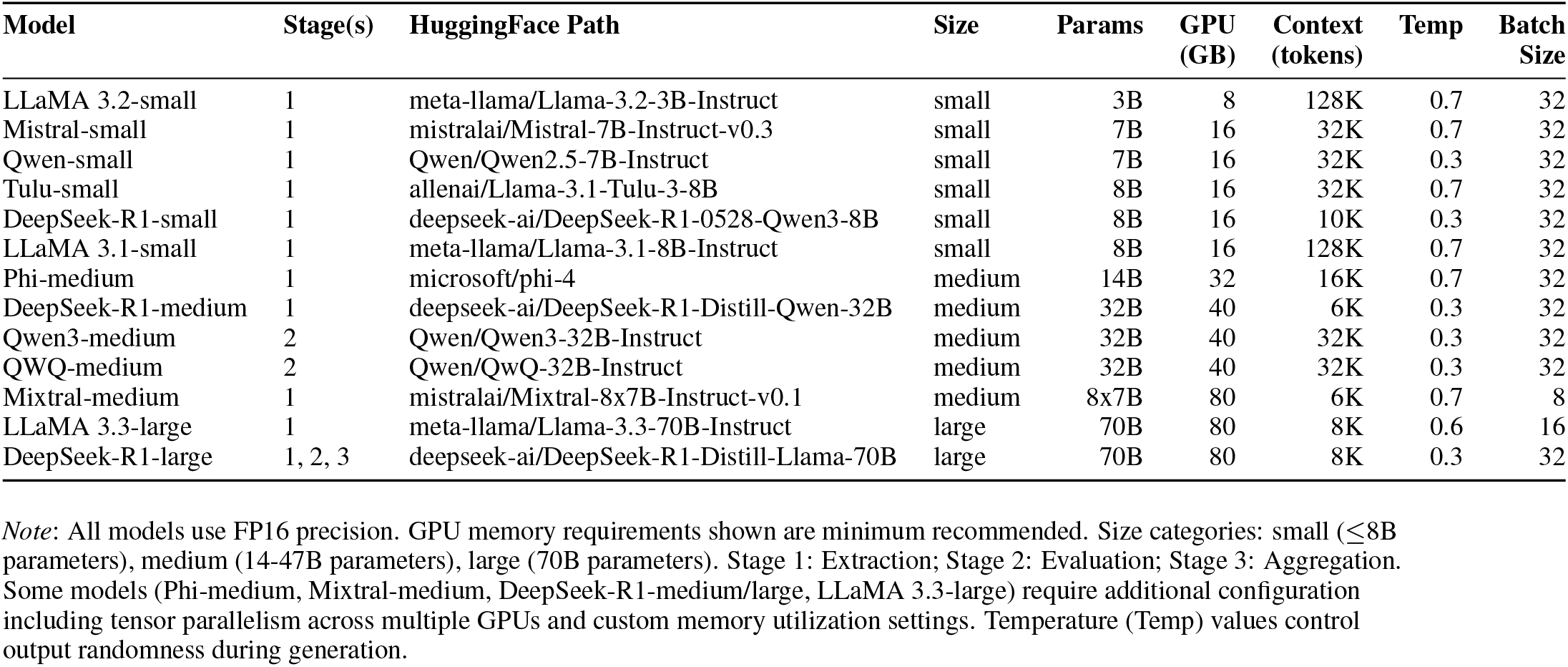
Model Zoo: Specifications of all LLMs used in the three-stage extraction framework.

| Model | Stage(s) | HuggingFace Path | Size | Params | GPU (GB) | Context (tokens) | Temp | Batch Size |
| --- | --- | --- | --- | --- | --- | --- | --- | --- |
| LLaMA 3.2-small | 1 | meta-llama/Llama-3.2-3B-Instruct | small | 3B | 8 | 128K | 0.7 | 32 |
| Mistral-small | 1 | mistralai/Mistral-7B-Instruct-v0.3 | small | 7B | 16 | 32K | 0.7 | 32 |
| Qwen-small | 1 | Qwen/Qwen2.5-7B-Instruct | small | 7B | 16 | 32K | 0.3 | 32 |
| Tulu-small | 1 | allenai/Llama-3.1-Tulu-3-8B | small | 8B | 16 | 32K | 0.7 | 32 |
| DeepSeek-R1-small | 1 | deepseek-ai/DeepSeek-R1-0528-Qwen3-8B | small | 8B | 16 | 10K | 0.3 | 32 |
| LLaMA 3.1-small | 1 | meta-llama/Llama-3.1-8B-Instruct | small | 8B | 16 | 128K | 0.7 | 32 |
| Phi-medium | 1 | microsoft/phi-4 | medium | 14B | 32 | 16K | 0.7 | 32 |
| DeepSeek-R1-medium | 1 | deepseek-ai/DeepSeek-R1-Distill-Qwen-32B | medium | 32B | 40 | 6K | 0.3 | 32 |
| Qwen3-medium | 2 | Qwen/Qwen3-32B-Instruct | medium | 32B | 40 | 32K | 0.3 | 32 |
| QWQ-medium | 2 | Qwen/QwQ-32B-Instruct | medium | 32B | 40 | 32K | 0.3 | 32 |
| Mixtral-medium | 1 | mistralai/Mixtral-8x7B-Instruct-v0.1 | medium | 8x7B | 80 | 6K | 0.7 | 8 |
| LLaMA 3.3-large | 1 | meta-llama/Llama-3.3-70B-Instruct | large | 70B | 80 | 8K | 0.6 | 16 |
| DeepSeek-R1-large | 1, 2, 3 | deepseek-ai/DeepSeek-R1-Distill-Llama-70B | large | 70B | 80 | 8K | 0.3 | 32 |

*Note:* All models use FP16 precision. GPU memory requirements shown are minimum recommended. Size categories: small ( $\leq 8$ B parameters), medium (14-47B parameters), large (70B parameters). Stage 1: Extraction; Stage 2: Evaluation; Stage 3: Aggregation. Some models (Phi-medium, Mixtral-medium, DeepSeek-R1-medium/large, LLaMA 3.3-large) require additional configuration including tensor parallelism across multiple GPUs and custom memory utilization settings. Temperature (Temp) values control output randomness during generation.

In the second stage, each extracted variable and its corresponding justification were evaluated by three separate LLMs acting as automated adjudicators: DeepSeek-R1:70B (DeepSeek-R1-large), Qwen3:32B (Qwen3-medium), and QWQ:32B (QWQ-medium). These evaluator models were chosen to provide diverse perspectives and improve robustness of the evaluation process. For every extraction, each evaluator was presented with the original pathology report text and the variable-specific justification provided by the LLM that produced the prediction. Based on internal consistency, textual evidence, and clinical plausibility, each evaluator assigned a binary score of 1 (correct) or 0 (incorrect) to each output. The combined evaluation scores from all three evaluators were used to identify high-confidence predictions and to inform downstream aggregation in the final stage. Inter-evaluator agreement was assessed using correlation analysis and intraclass correlation coefficients.

In the third stage, a reasoning-based aggregation process was used to determine final consensus values for each variable. DeepSeek-R1-large was used as the aggregation model, prompted to adjudicate among the outputs of all eleven LLMs using the combined evaluation scores from all three evaluators in Stage 2. For each variable, the aggregation model synthesized the predictions, justifications, and multi-evaluator scores to select a single consensus value. The model prioritized outputs with both inter-model agreement and high evaluation scores across multiple evaluators, while discounting hallucinated or unsupported responses. Final outputs were formatted as structured JSON objects that included not only the selected values but also concise rationales explaining the reasoning behind each choice. These justifications were designed to enable human-in-the-loop review and support traceability in clinical workflows.

### 2.2 Expert Review and Clinical Validation

To evaluate clinical validity and real-world usability, a subset of final outputs from the aggregation stage was independently reviewed by board-certified (or holding equivalent qualification) pathologists. Reviewers were presented with the original pathology report alongside the model-extracted values (from stage 3) and corresponding justifications. Each variable was scored on a three-point scale: 1 (correct), 0.5 (partially correct), or 0 (incorrect), based on consistency with the report content and accepted clinical interpretation. In addition to assigning scores, reviewers provided qualitative feedback to identify common error types, such as omission of laterality, stage misclassification due to missing contextual cues, and under-classification of tumor grade. These expert annotations helped validate model performance in nuanced clinical scenarios and informed recommendations for refining prompts, improving reasoning trace quality, and flagging low-confidence outputs for human review.

### 2.3 Datasets

The framework was evaluated using two datasets of surgical pathology reports: one derived from TCGA and the other from patient records at Moffitt Cancer Center. The TCGA dataset was sourced from the OCR-extracted corpus released by Kefeli and Tatonetti [33], from which we randomly selected 6,100 reports spanning ten organ sites: kidney (858), breast (820), lung (750), uterus (639), brain (620), prostate (472), liver (421), bladder (413), cervix (309), and pancreas (139). This expanded dataset enabled evaluation across a broader spectrum of cancer types and reporting styles. A subset of 138 reports from five organs (bladder, prostate, liver, cervix, and kidney) was manually reviewed by three board-certified (or having equivalent qualification) pathologists (414 evaluations in total) to support expert evaluation.

The Moffitt dataset consisted of 510 de-identified surgical pathology reports from patients treated between 2009 and 2018. All reports were linked to institutional cancer registry records and included three cancer types: breast (170), brain or central nervous system (170), and lung (170). For these reports, both standard diagnostic variables and organ-specific biomarkers were extracted. For expert review, 30 reports were randomly selected from each organ group (90 total) and independently evaluated by five board-certified (or having equivalent qualification) pathologists, including specialists in each organ system (DSV, FK, ZGO for Moffitt; AK, EU, WSC for TCGA). All reports in both datasets were converted into structured JSON format containing a unique identifier, organ label, and full free-text report, allowing for consistent preprocessing, prompting, and scoring across the three-stage framework.

### 2.4 Statistical Analysis

Statistical analyses were conducted separately for the automated model evaluations and the expert-reviewed subsets. For each dataset, structured records containing model identity, variable type, organ site, evaluator identity, and extraction score were compiled for analysis. One-way ANOVA was used to compare average extraction accuracy across the eleven LLMs and to assess differences between the three evaluator models. Two-way ANOVA was used to assess interactions between model type and diagnostic variable, as well as evaluator and model interactions. A three-way ANOVA was used to examine joint effects of model, variable, and organ site, including all pairwise and three-way interaction terms. Inter-evaluator agreement was assessed using Pearson correlations and intraclass correlation coefficients (ICC), with ICC values interpreted as: *<* 0.5 (poor), 0.5 − 0.75 (moderate), 0.75 − 0.9 (good), and *>* 0.9 (excellent agreement). When statistically significant main or interaction effects were observed, Tukey’s Honestly Significant Difference (HSD) was used to identify pairwise differences between models or organs while controlling the family-wise error rate. For the biomarker dataset, additional analyses were performed to evaluate organ-specific biomarker extraction performance.

For the expert-reviewed subsets, the pathologist-assigned accuracy scores were analyzed using two-way ANOVA with fixed effects for variable and organ. This analysis was conducted separately for the TCGA and Moffitt datasets. As with the automated analysis, significant differences were followed by Tukey’s HSD tests to assess specific pairwise contrasts. All statistical tests were performed using the statsmodels package in Python (v0.14.0), with significance defined as (*α* = 0.05).

## 3 Results

The performance of eleven locally deployed LLMs was evaluated in extracting six clinically relevant variables, site, laterality, histology, stage, grade, and behavior, from unstructured surgical pathology reports. The evaluation was conducted across two distinct datasets: publicly available pathology reports from TCGA and patient reports from Moffitt Cancer Center. For each dataset, a two-tiered assessment strategy was applied: automated scoring by a reference LLM based on the extracted values and justifications, and expert review by board-certified or equivalent pathologists on a randomly selected subset. Statistical comparisons were performed to assess variability in extraction accuracy across models, variables, and organ types. Results are presented in two parts: TCGA and Moffitt. Each part is organized by evaluation method: automated scoring followed by expert validation.

### 3.1 Expert Evaluation: Combined TCGA and Moffitt Datasets

Figure 2 presents comprehensive expert evaluation results from both TCGA and Moffitt datasets. Panel (A) shows expert evaluation results from 138 randomly selected subset of TCGA pathology reports spanning five organ types (bladder, prostate, liver, cervix, and kidney), with each variable scored by board-certified pathologists on a three-point scale (1=correct, 0.5=partially correct, 0=incorrect). TCGA reports demonstrated high overall accuracy, with behavior (100%), histology (99%), grade (97%), and site (95%) achieving the strong performance, while stage (88%) and laterality (87%) showed more variability. Panel (B) presents expert evaluation of 180 randomly selected subset of Moffitt reports covering brain, breast, and lung cancers. Moffitt reports showed comparably high accuracy, with behavior (99%), histology and laterality (96%), grade (93%), and site (91%). Panel (C) provides a direct comparison of performance on the three organs common to both datasets (brain, breast, and lung), revealing consistent extraction accuracy across institutions with slight variations in organ-specific performance patterns. The comparative analysis highlights that institutional reporting styles had minimal impact on extraction accuracy for standard diagnostic variables.

**Figure 2:**
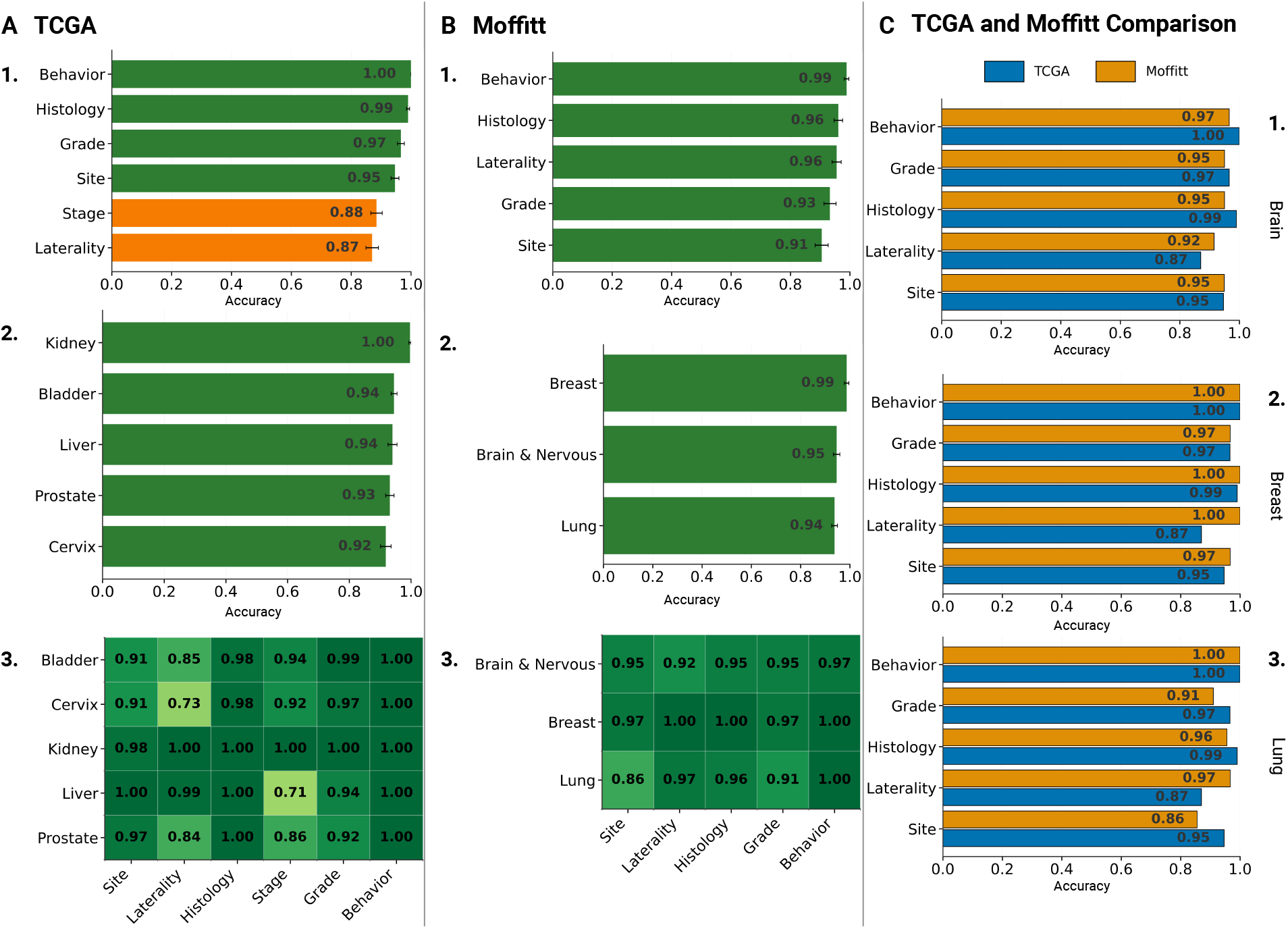
Expert evaluation of structured variable extraction across TCGA and Moffitt datasets. This figure presents comprehensive expert validation results from both institutional and public datasets. **Panel (A)** displays TCGA expert evaluation results from 138 randomly selected pathology reports spanning five organ types (bladder, prostate, liver, cervix, and kidney), with performance metrics for six diagnostic variables scored by board-certified pathologists. The heatmap shows variable-organ interactions with behavior achieving perfect accuracy (100%) and laterality showing the most variability (84.8%). **Panel (B)** presents Moffitt expert evaluation from 180 reports covering brain, breast, and lung cancers, demonstrating comparably high accuracy with behavior (99%) and histology (96%) performing strongly. **Panel (C)** provides a direct comparative analysis of the three organs common to both datasets (brain, breast, and lung), revealing consistent extraction accuracy across institutions despite different reporting styles. The comparative visualization highlights minimal impact of institutional documentation practices on extraction performance for standard diagnostic variables.

Statistical analysis of expert evaluations revealed significant effects of both variable type and organ. For TCGA, two-way ANOVA showed main effects for variable (*F* (5, 1195) = 16.95, *p <* 0.0001) and organ (*F* (4, 1195) = 3.94, *p* = 0.0035), with a significant interaction (*F* (20, 1195) = 4.46, *p <* 0.0001). Post-hoc tests identified that laterality extraction was significantly more accurate in kidney and liver compared to cervix (*p* = 0.005 and *p* = 0.0025), while stage showed lower accuracy in liver than bladder, cervix, and kidney (all *p <* 0.01). For Moffitt, extraction accuracy was affected by variable type (*F* (5, 997) = 4.05, *p* = 0.003) and the variable-organ interaction (*F* (10, 997) = 3.05, *p <* 0.001), with site extraction more accurate in brain and breast than lung (*p* = 0.017 and *p* = 0.039).

### 3.2 TCGA Dataset: Automated Evaluation

Figure 3 presents a comprehensive analysis of automated evaluation results from the TCGA dataset using three independent evaluator models. Panel (A) displays the average evaluation scores assigned by the three evaluators (DeepSeek-R1-large, Qwen3-32B, and QWQ-32B) to each of the eleven extraction models. The mean accuracy across all evaluations was 84.9%±7.3%, with top-performing models including Phi-4-medium, DeepSeek-R1-70b and 32b, and LLaMA 3.3 consistently achieving highest scores, while Mixtral-medium, Mistral-small, DeepSeek-R1-small, and LLaMA 3.2 showed lower performance across evaluators. Panel (B) illustrates the concordance between evaluators through correlation analysis, revealing high inter-evaluator agreement with Pearson correlations exceeding 0.93 (DeepSeek-R1-large vs Qwen3-32B: r=0.943; DeepSeek-R1-large vs QWQ-32B: r=0.939; Qwen3-32B vs QWQ-32B: r=0.955), *p <* 0.05, confirming the robustness of the multi-evaluator approach.

**Figure 3:**
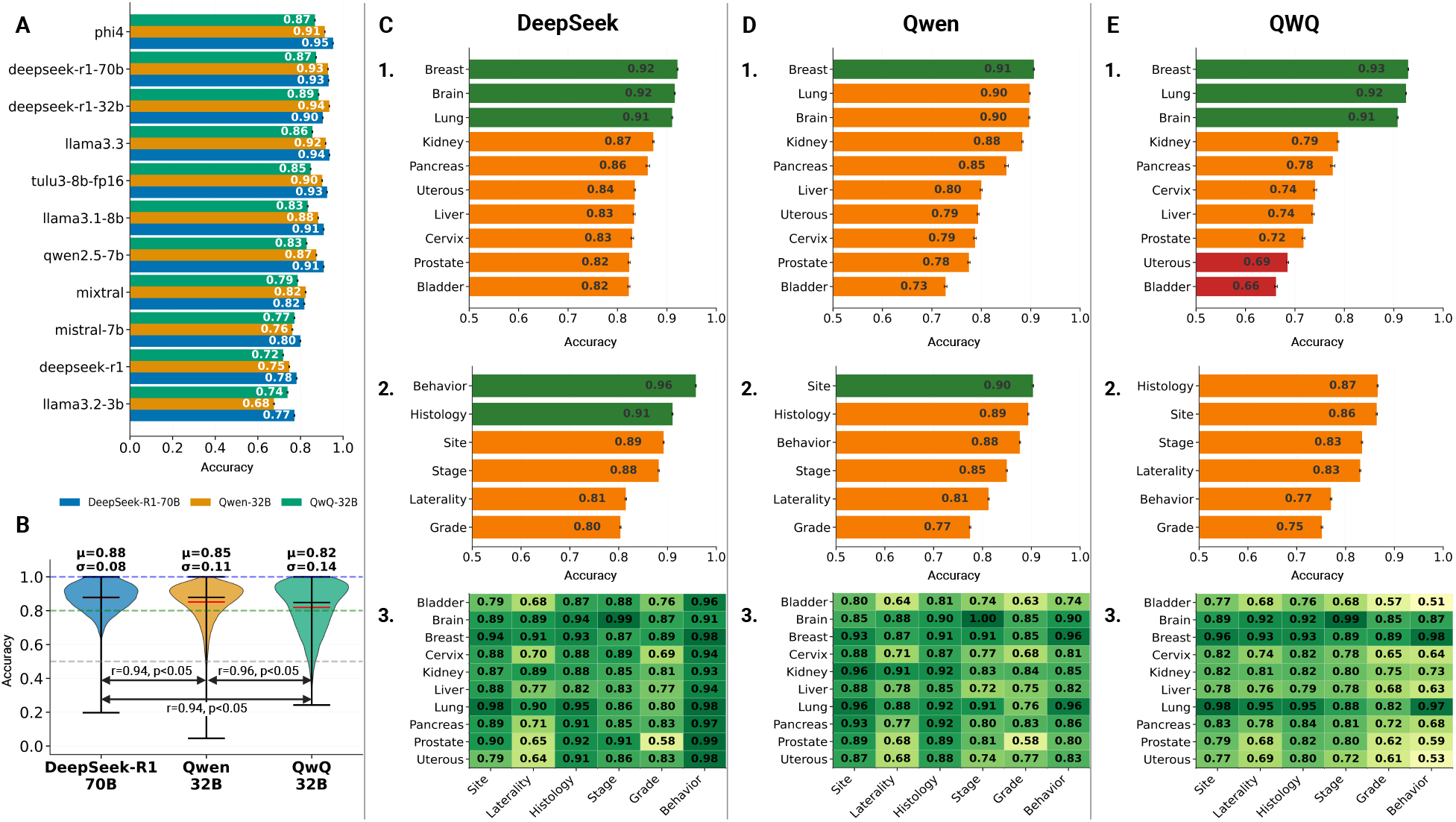
Multi-evaluator automated assessment of extraction performance on the TCGA dataset. This figure presents comprehensive automated evaluation results from 6,100 TCGA pathology reports across ten organ types using three independent evaluator models. **Panel (A)** displays average evaluation scores assigned by DeepSeek-R1-large, Qwen3-32B, and QWQ-32B to each of the eleven extraction models, with mean accuracy of 84.9% ± 7.3%. Top performers (Phi-4-medium, DeepSeek-R1-70b and 32b, LLaMA 3.3) consistently achieved >86% accuracy. **Panel (B)** illustrates evaluator concordance through correlation matrices, revealing high inter-evaluator agreement (*r >* 0.94 for all pairs), confirming robustness of the multi-evaluator approach. **Panels (C-E)** provide evaluator-specific analyses with three subpanels each: (C) DeepSeek-R1-large showing highest organ accuracy (breast, brain, and lung), variable accuracy (behavior and histology best), and a comprehensive heatmap; (D) Qwen3-32B demonstrating consistent patterns with slight model ranking variations; (E) QWQ-32B displaying highest concordance with DeepSeek-R1-large while maintaining unique assessment characteristics. The multi-evaluator framework revealed complementary perspectives particularly valuable for ambiguous extractions.

Panels (C), (D), and (E) provide detailed evaluator-specific analyses. Panel (C) focuses on DeepSeek-R1-large’s evaluations, showing organ-wise accuracy (highest for kidney and pancreas), variable-wise accuracy (behavior and histology performing best), and a comprehensive heatmap revealing model-variable-organ interactions. Panel (D) presents similar analyses for Qwen3-32B, demonstrating consistent patterns with slight variations in model rankings. Panel (E) displays QWQ-32B’s evaluations, which showed the highest concordance with DeepSeek-R1-large while maintaining unique assessment characteristics. The multi-evaluator approach revealed that while overall agreement was high, each evaluator contributed complementary perspectives, particularly in edge cases involving ambiguous or complex extractions.

Statistical analysis confirmed significant performance differences across all factors. A one-way ANOVA revealed overall differences in extraction accuracy across the eleven LLMs (*F* = 1716.82, *p <* 0.001). A two-way ANOVA showed significant main effects for model (*F* = 1770.86, *p <* 0.001) and variable (*F* = 3236.68, *p <* 0.001), along with a strong interaction (*F* = 175.86, *p <* 0.001), indicating that no single model performed uniformly across all variables. A three-way ANOVA incorporating organ type revealed additional variation, with significant effects for organ (*F* = 1946.43, *p <* 0.001) and multiple interaction terms (model × organ: *F* = 268.13, *p <* 0.001; variable × organ: *F* = 375.83, *p <* 0.001; model × variable × organ: *F* = 24.74, *p <* 0.001), highlighting the influence of organ-specific language patterns on model accuracy. Evaluator comparison revealed significant differences (*F* = 2633.79, *p <* 0.001) despite high inter-evaluator agreement. Post-hoc Tukey HSD tests identified that Mixtral-medium underperformed significantly on histology, laterality, and stage compared to higher-performing models like DeepSeek-R1-large and Phi-4-medium.

### 3.3 Moffitt Dataset: Automated Evaluation

Figure 4 presents the automated evaluation results from 510 Moffitt pathology reports using the same three-evaluator framework applied to the TCGA dataset. Panel (A) shows the average scores assigned by the three evaluators to each extraction model, with mean accuracy of 88.2%±7.2%, demonstrating higher overall performance than TCGA despite the institutional-specific reporting styles. Top-performing models (Phi-4-medium, Tulu3-small, DeepSeek-R1-32b, LLaMA 3.3, DeepSeek-R1-70b, and LLaMA 3.1) achieved consistently high scores across all evaluators, while Mixtral-medium, DeepSeek-R1-small, LLaMA-small, and Mistral-small showed reduced performance particularly on grade and histology extraction. Panel (B) demonstrates evaluator concordance with high correlations (DeepSeek-R1-large vs Qwen3-32B: r=0.934; DeepSeek-R1-large vs QWQ-32B: r=0.981; Qwen3-32B vs QWQ-32B: r=0.939), *p <* 0.05, confirming consistency across evaluators despite the different institutional context.

**Figure 4:**
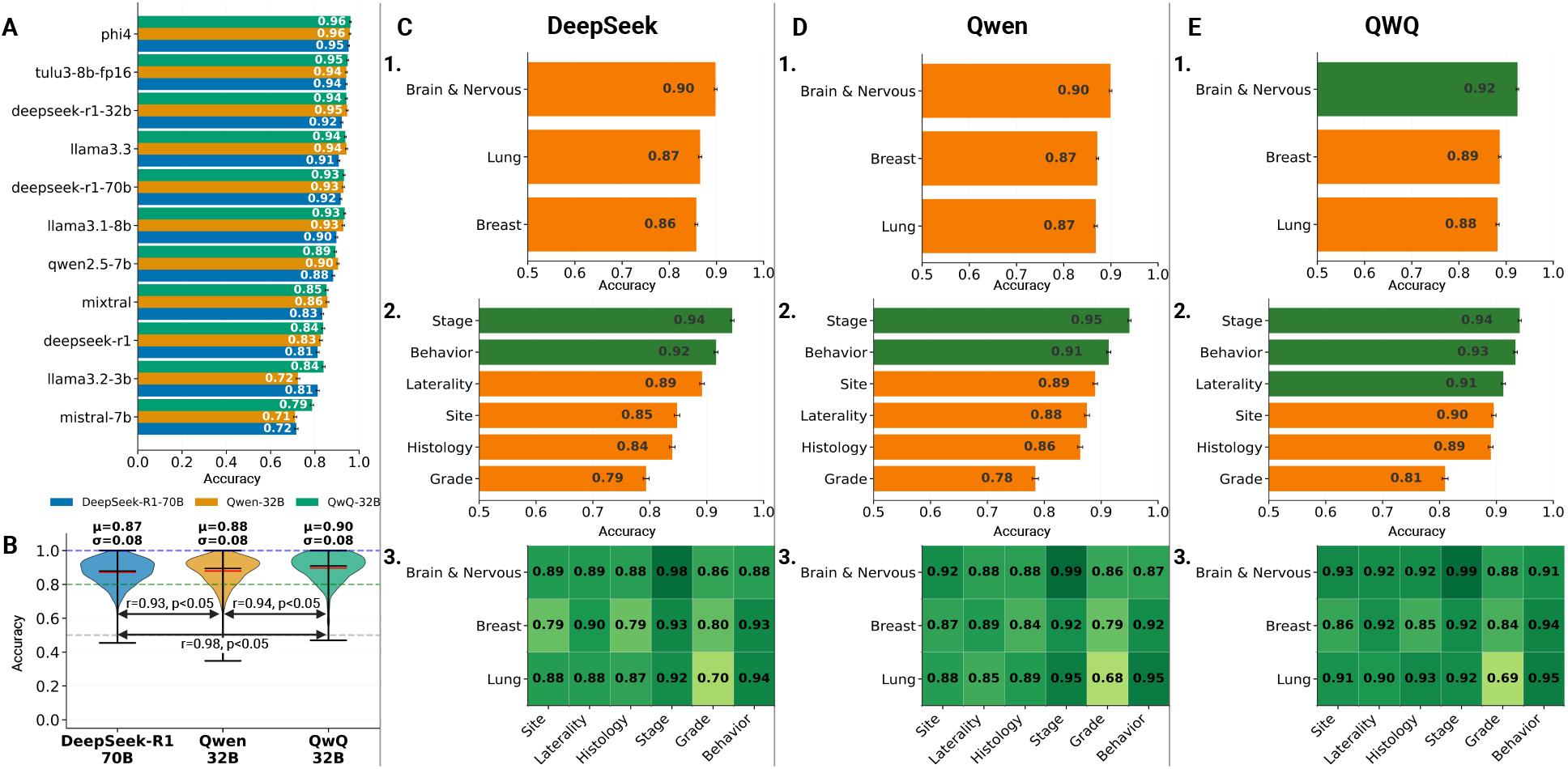
Multi-evaluator automated assessment of extraction performance on the Moffitt institutional dataset. This figure presents automated evaluation results from 510 Moffitt pathology reports (brain, breast, lung) using the three-evaluator framework. **Panel (A)** shows average scores from three evaluators for eleven extraction models, achieving 88.2% ± 7.2% mean accuracy—higher than TCGA despite institutional-specific reporting. Top performers (Phi-4-medium, Tulu3-small, DeepSeek-R1-32b, LLaMA 3.3, DeepSeek-R1-70b, and LLaMA 3.1) maintained consistency across evaluators. **Panel (B)** demonstrates evaluator concordance with high correlations (DeepSeek-R1-large vs Qwen3-32B: r=0.93; DeepSeek-R1-large vs QWQ-32B: r=0.98; Qwen3-32B vs QWQ-32B: r=0.94), *p <* 0.05. **Panels (C-E)** provide evaluator-specific analyses: (C) DeepSeek-R1-large revealing uniform organ performance and strong stage/behavior extraction (94%, 92%); (D) Qwen3-32B showing similar patterns with variations in biomarker-related rankings; (E) QWQ-32B displaying highest correlation with DeepSeek-R1-large (r=0.98). The uniform cross-organ performance, contrasting with TCGA variability, suggests institutional documentation practices may enhance extraction consistency.

Panels (C), (D), and (E) present individual evaluator analyses. Panel (C) shows DeepSeek-R1-large’s assessments with organ-specific accuracy revealing uniform performance across brain, breast, and lung reports, variable-specific accuracy highlighting strong performance on stage (94%) and behavior (92%), and a heatmap showing model-variable interactions. Panel (D) displays Qwen3-32B’s evaluations with similar patterns but slight variations in model rankings, particularly for biomarker-related extractions. Panel (E) presents QWQ-32B’s results, which showed the highest correlation with DeepSeek-R1-large (r=0.981) while maintaining independent assessment characteristics. The uniform performance across organs in the Moffitt dataset, compared to greater variability in TCGA, suggests that institutional-specific documentation practices may actually enhance extraction consistency.

Statistical analysis confirmed significant differences in extraction accuracy across models and variables. A one-way ANOVA revealed a significant main effect of model (*F* = 477.31, *p <* 0.001), indicating performance heterogeneity among the eleven LLMs. Evaluator comparison showed significant differences (*F* = 52.93, *p <* 0.001), though inter-evaluator agreement remained high (correlations: DeepSeek-R1-large vs Qwen3-32B: r=0.934; DeepSeek-R1-large vs QWQ-32B: r=0.981; Qwen3-32B vs QWQ-32B: r=0.939). Two-way ANOVA showed significant main effects for both model and variable, along with a model × variable interaction, confirming that accuracy depended on the specific variable being extracted. When organ was added as a third factor, a main effect for organ was observed (*F* = 7.84, *p <* 0.001), as well as a significant interaction between variable and organ (*F* = 17.69, *p <* 0.001). However, no significant model × organ or three-way interaction effects were found, suggesting greater consistency in model performance across organs compared to the TCGA cohort. Post-hoc Tukey HSD tests showed that Mixtral-medium underperformed significantly on histology, laterality, and stage compared to DeepSeek and other top models, while behavior and site remained consistently well-predicted across models.

### 3.4 Moffitt Biomarker Dataset: Automated Evaluation

Figure 5 presents comprehensive biomarker extraction results from the same 510 Moffitt pathology reports, with organ-specific analyses across three cancer types. Overall accuracy for biomarker extraction was 70.6%±8.1%, reflecting the increased complexity of identifying molecular markers from narrative text compared to standard diagnostic variables. Panel (A) focuses on brain tumor biomarkers, with three subpanels showing: (1) average biomarker extraction accuracy across models, where TP53 achieved 85%, followed by 1p/19q codeletion (77%), ATRX (73%), MGMT methylation (75%), and IDH1 (71%); (2) evaluator LLMs’ comparison; and (3) a heatmap revealing model-specific performance patterns across evaluator LLMs. IDH2 mutation status, often reported using various nomenclatures, achieved moderate average accuracy (67%) across all models.

**Figure 5:**
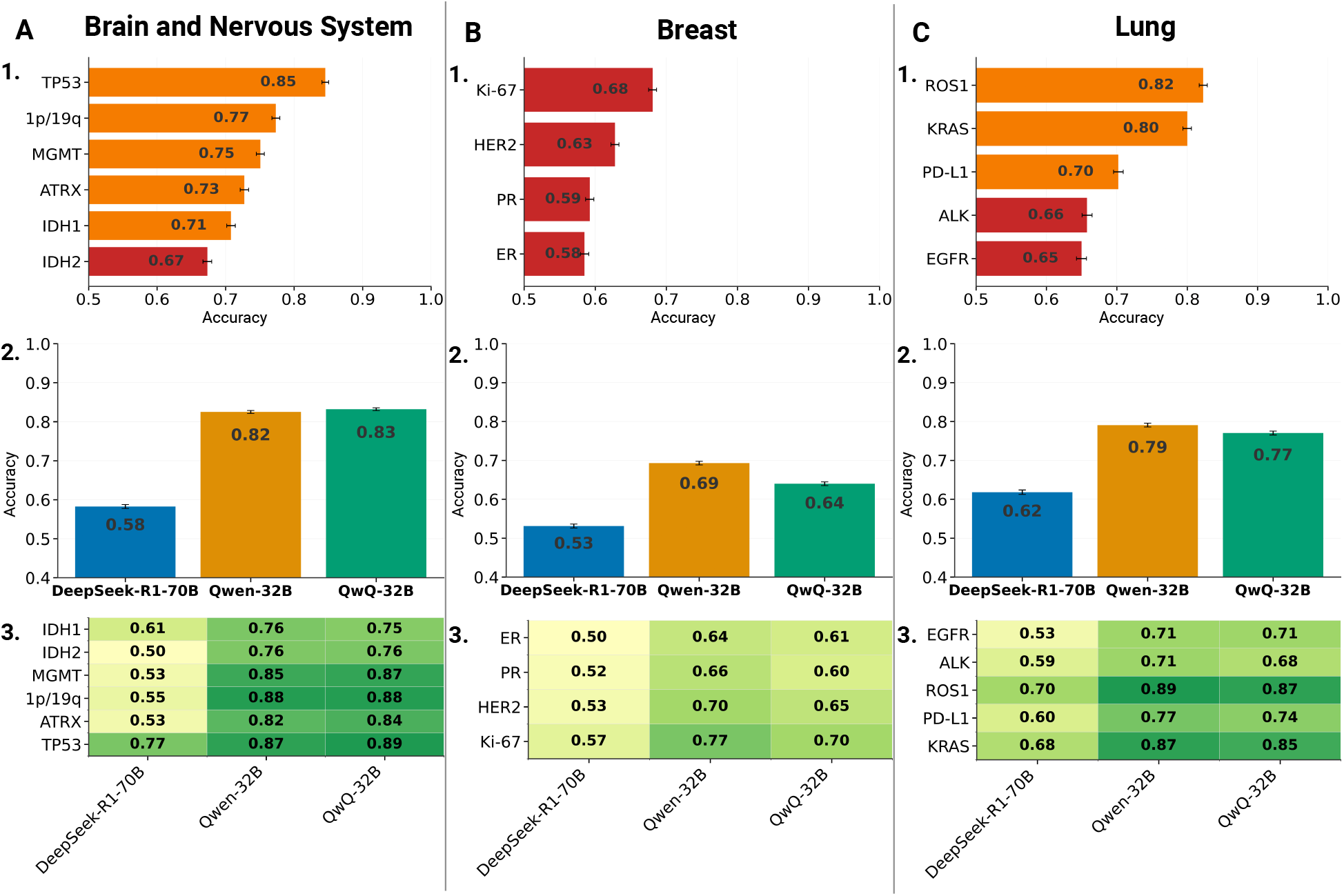
Organ-specific biomarker extraction performance from Moffitt pathology reports. This figure presents comprehensive biomarker extraction results from 510 Moffitt reports with overall accuracy of 70.6% ± 8.1%, reflecting increased complexity of molecular marker identification. **Panel (A) - Brain Biomarkers:** Three subpanels showing (1) average accuracy with TP53 achieving 85% accuracy, followed by 1p/19q codeletion (77%), ATRX (73%), MGMT (75%) and IDH1 (71%); (2) evaluator concordance demonstrating high agreement despite neurological biomarker complexity; (3) model-biomarker heatmap revealing LLMs’ superiority on complex markers. **Panel (B) - Breast Biomarkers:** Subpanels displaying (1) hormone receptor accuracy: Ki-67 (68%), HER2 (63%), PR (59%), ER (58%); (2) lower evaluator agreement suggesting greater interpretative complexity; (3) heatmap showing extraction challenges from complex reporting formats. **Panel (C) - Lung Biomarkers:** Subpanels presenting (1) strong performance for ROS1 (82%) and KRAS (80%), moderate for PD-L1 (70%), ALK (66%), and EGFR (65.0%); (2) high evaluator concordance (r > 0.89) despite biomarker diversity; (3) model-specific molecular marker detection strengths. Statistical analysis confirmed significant model effects (*F* = 538.00, *p <* 0.001) with robust multi-evaluator agreement.

**Figure 6:**
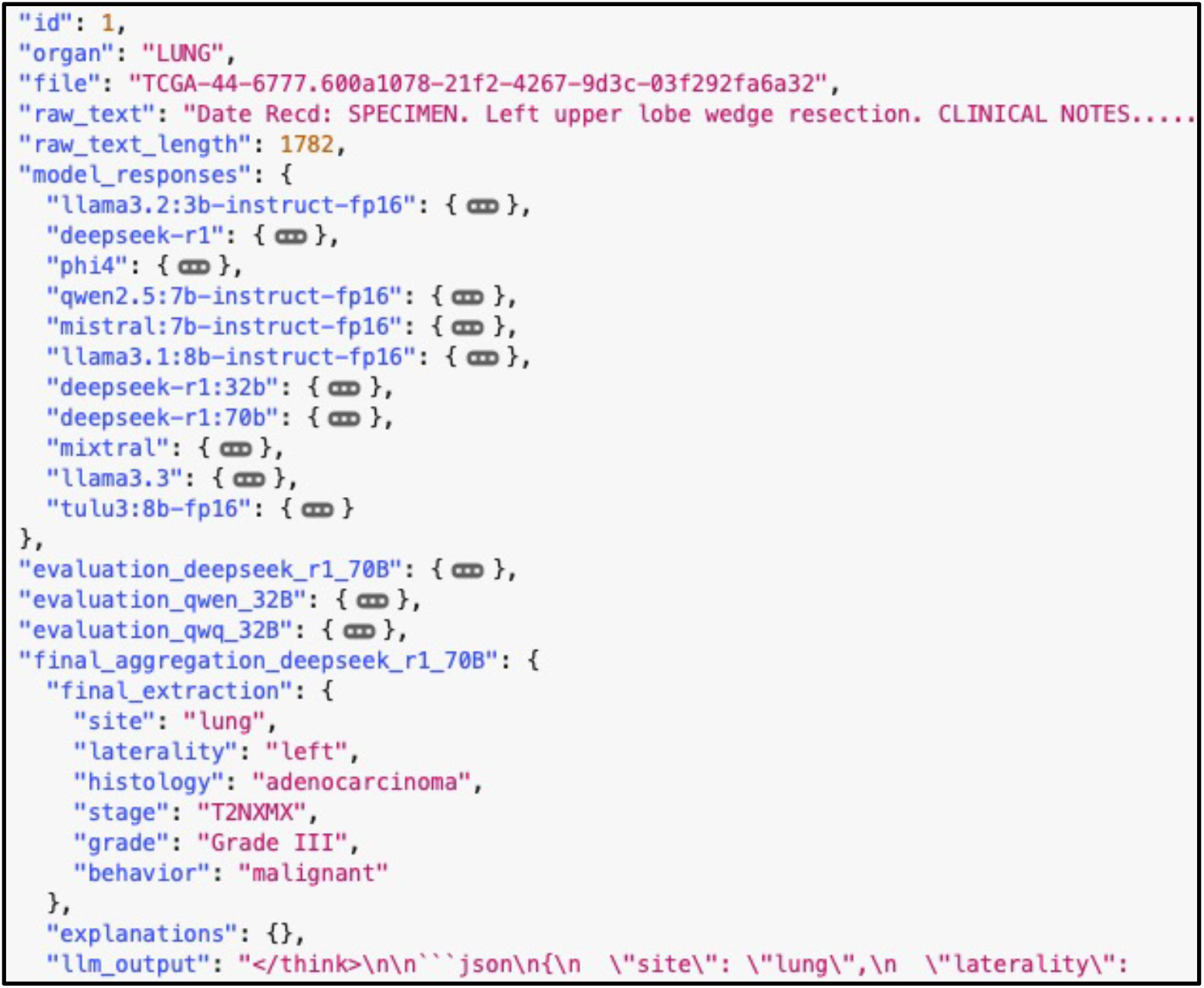
Example JSON record produced by the three-stage clinical extraction framework. The entry corresponds to a TCGA lung pathology note (“Left upper lobe wedge resection”) and shows the complete artifacts for one case. Top-level metadata include the internal id, organ/site (“LUNG”), file handle, the original note and raw_text_length. Under model_responses are the Stage-1 extractor candidates generated by each model in the Model Zoo (e.g., llama3.2:3b-instruct-fp16, Tulu3-small-fp16, DeepSeek-R1-medium/large, phi4, qwen2.5:7b, mistral:7b, Mixtral-medium, llama3.1:8b, llama3.3), each producing a schema-constrained JSON for the target variables. The evaluation_* keys store Stage-2 evaluator judgments from Qwen-32B, QwQ-32B, and DeepSeek-R1-70B over those candidates. The final_aggregation_deepseek_r1_70B block records the Stage-3 aggregator’s decision and the canonical final_extraction, here resolving to: site = lung, laterality = left, histology = adenocarcinoma, stage = TNM string, grade = Grade III, and behavior = malignant. This record illustrates how the system keeps raw candidates, evaluator signals, and the aggregated, ready-to-analyze output for auditability and downstream analysis.

**Figure 7:**
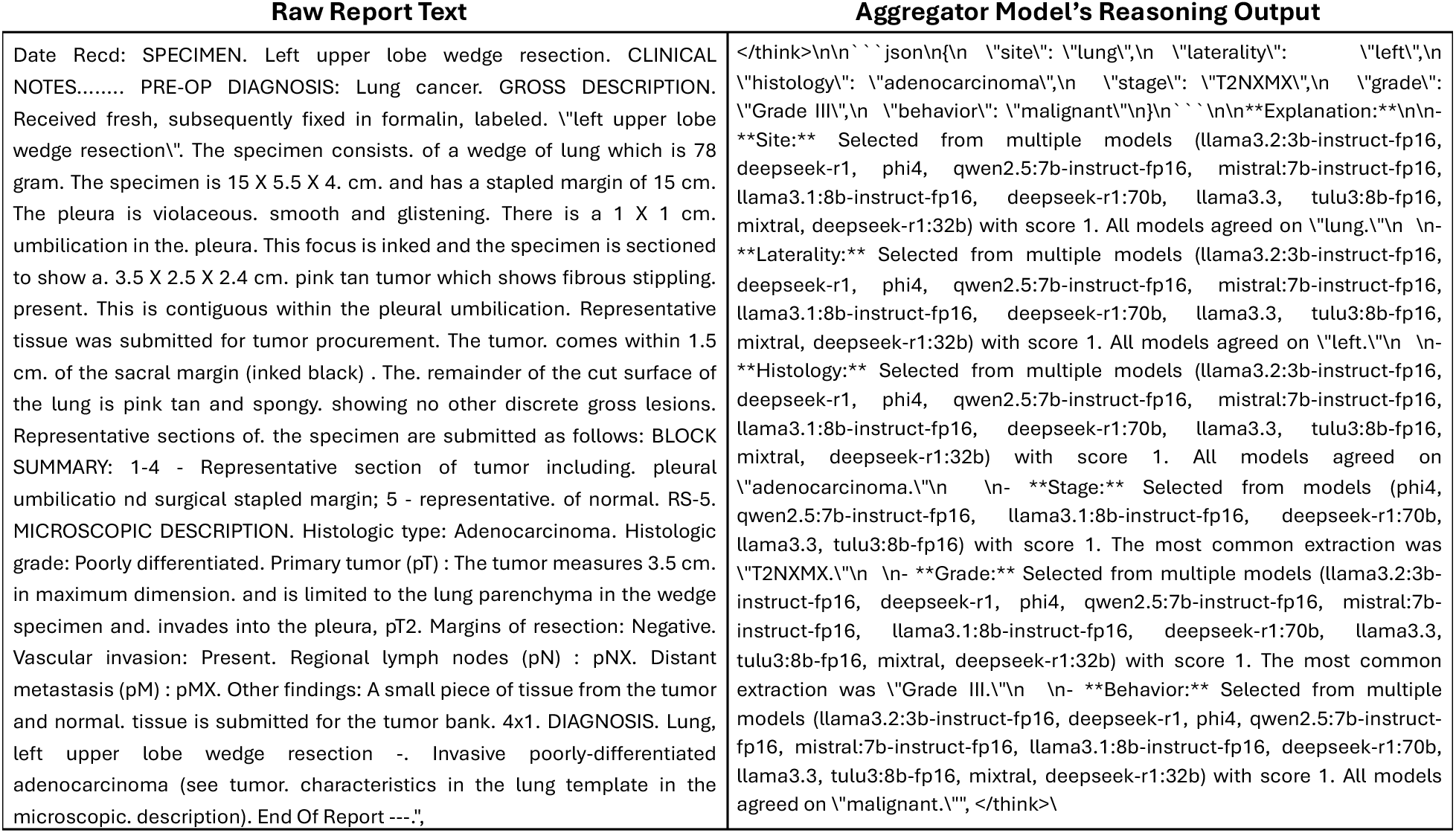
Example of end-to-end artifacts from the three-stage clinical information-extraction framework. (Left): De-identified raw pathology report text for a TCGA lung resection, including gross, microscopic, and synoptic elements. (Right): The aggregator model’s “thinking extract,” which logs the schema-constrained final extraction JSON (site, laterality, histology, stage, grade, behavior) together with a concise rationale summarizing cross-model consensus from the Stage-1 extractor ensemble. In this case, the aggregator resolves to site = lung, laterality = left, histology = adenocarcinoma, stage = T2NXMX, grade = Grade III, and behavior = malignant, noting unanimous or majority agreement across candidate outputs. These audit trails illustrate how raw narrative evidence is transformed into structured variables while preserving provenance for quality assurance and error analysis.

**Figure 8:**
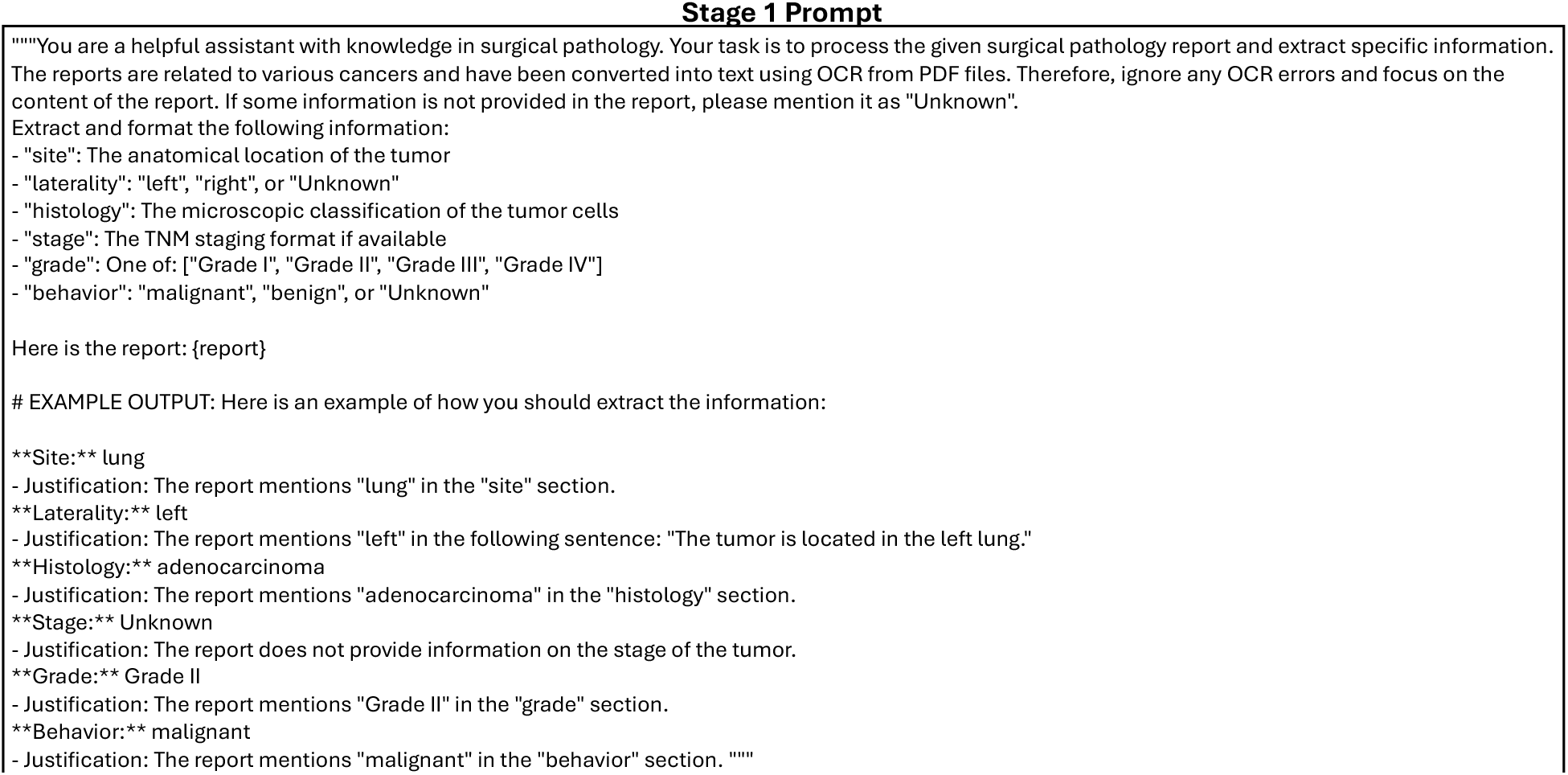
Stage 1 - Extraction Prompt. Complete instruction shown to models for schema-constrained clinical information extraction from raw surgical pathology text. The prompt specifies the six target fields (site, laterality, histology, stage, grade, behavior), asks models to ignore OCR artifacts, enforces allowed surface forms (e.g., “Grade I-IV”, “malignant/benign/Unknown”), and requires a brief justification per field with an example to standardize outputs.

**Figure 9:**
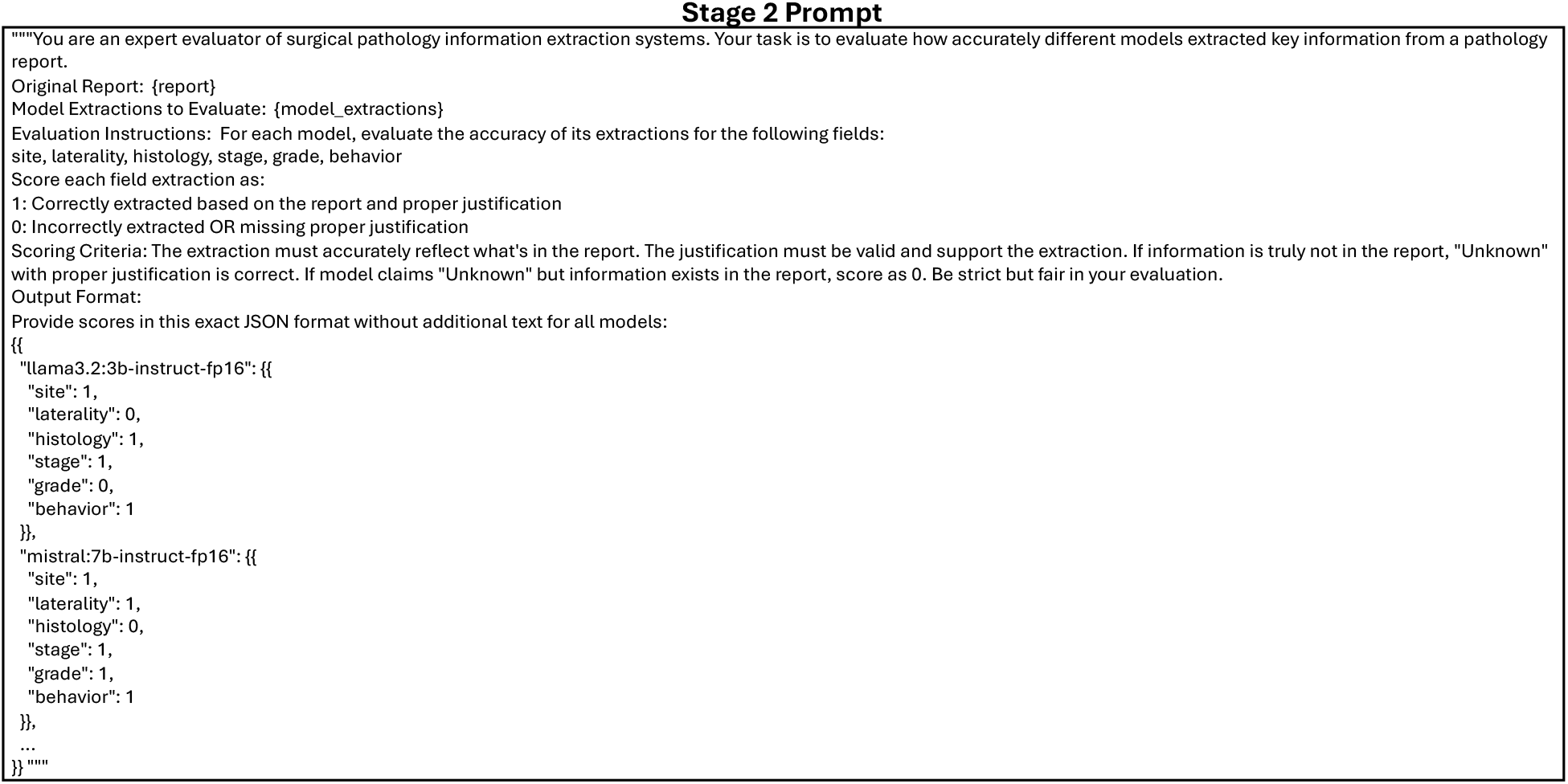
Stage 2 - Evaluator Prompt. Instruction given to automated evaluators that score each candidate extraction against the source report. For every model and field, the evaluator returns strict binary accuracy (1/0) contingent on both content correctness and a valid justification, in a JSON-only format. “Unknown” is correct only when the report truly lacks the information.

**Figure 10:**
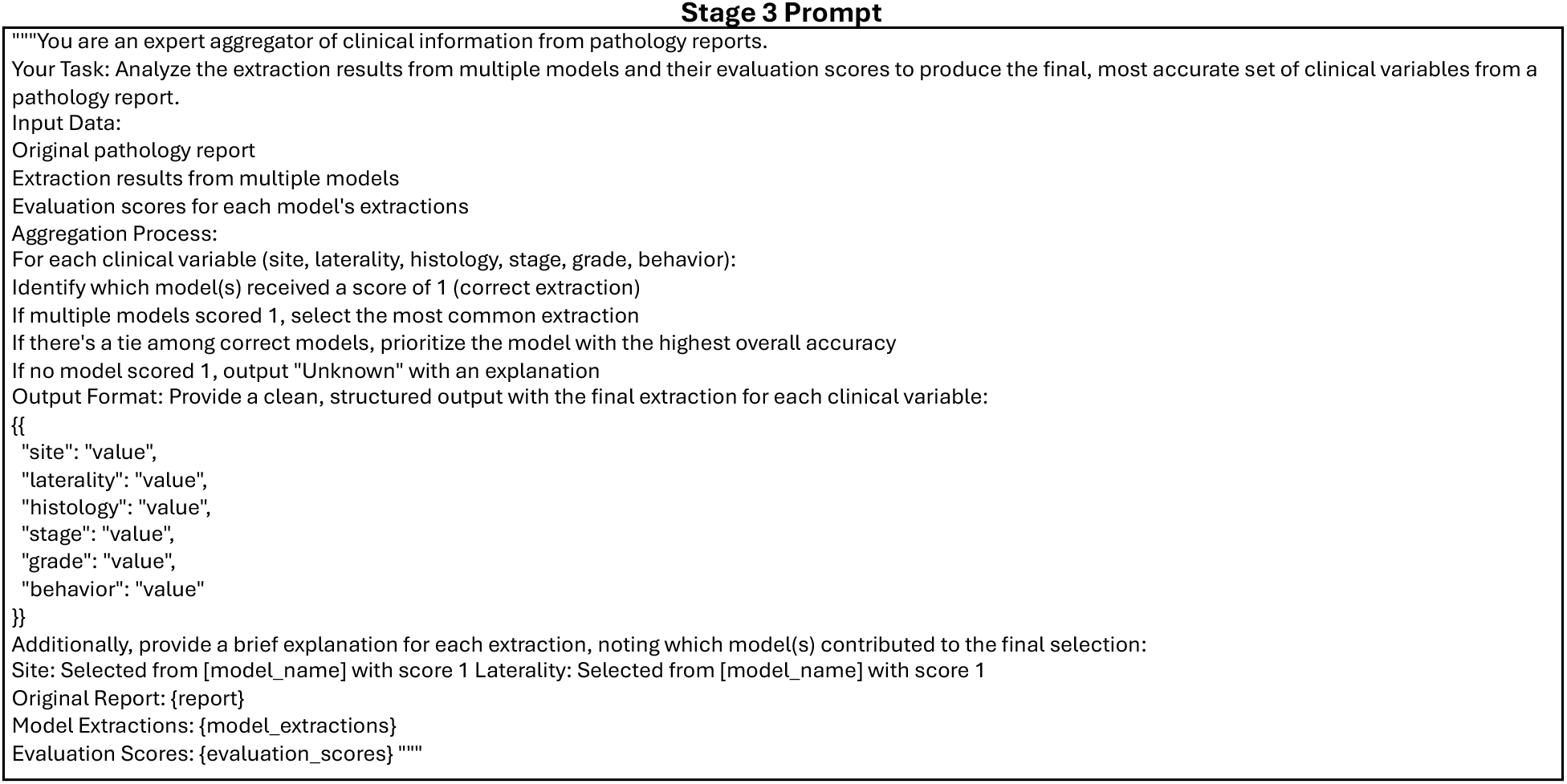
Stage 3 - Aggregator Prompt. Meta-prompt that ingests the original report, all candidate extractions, and evaluator scores to select the final labels. The procedure chooses the most common extraction among models that scored 1, breaks ties by the source model’s overall accuracy, and outputs “Unknown” with an explanation if no candidate is correct. The aggregator also emits brief, per-field rationales noting which model(s) contributed to each final selection.

Panel (B) examines breast cancer biomarkers, with subpanels displaying: (1) average accuracy for hormone receptors and proliferation markers, showing Ki-67 (68%), HER2 (63%), PR (59%), and ER (58%); (2) evaluator analysis showing slightly lower agreement than for brain markers especially between Qwen and QwQ, suggesting greater interpretative complexity in breast biomarker assessment; and (3) a model-biomarker heatmap revealing particular challenges with receptor status extraction due to complex reporting formats and multiple specimen types.

Panel (C) analyzes lung cancer biomarkers, with subpanels presenting: (1) average extraction accuracy highlighting strong performance for ROS1 (82%) and KRAS (80%), with PD-L1 expression at 70% and moderate performance for ALK (66%) and EGFR (65%); (2) evaluator LLMs’ comparison, where statistical analysis confirmed significant model effects (*F* = 538.00, *p <* 0.001) and robust multi-evaluator agreement across all biomarker categories; (3) and a comprehensive heatmap showing model-specific strengths in molecular marker detection.

## 4 Discussion

This study demonstrates that reasoning-based LLMs, when locally deployed and guided through structured prompting, can accurately and transparently extract both standard diagnostic variables and organ-specific biomarkers from free-text surgical pathology reports. Using a three-stage framework with multi-evaluator consensus applied to over 6,600 reports from ten cancer sites, including a large institutional cohort from Moffitt Cancer Center, we achieved high overall accuracy in extracting six clinically relevant fields (84.9% for TCGA, 88.2% for Moffitt) and moderate accuracy for biomarkers (70.6%). Automated evaluation using three independent LLM adjudicators (DeepSeek-R1-large, Qwen3-32B, and QWQ-32B) with high inter-evaluator agreement (*r >* 0.89) identified consistent accuracy trends, with histology, site, and behavior emerging as the most reliably extracted variables. These findings were corroborated by expert review from board-certified pathologists (or pathologists holding specialist qualifications outside the US), who confirmed high fidelity across most variables and noted specific challenges for laterality and stage in select organ contexts. Together, these results support the technical robustness, clinical validity, and real-world adaptability of our consensus-based extraction framework for routine and research applications in surgical pathology.

Differences in extraction accuracy across diagnostic variables reflect both model-level challenges and variation in how clinical information is recorded across pathology reports. Variables such as behavior and histology were consistently extracted with high accuracy, likely due to their relatively standardized representation and clearer phrasing in most documents. In contrast, stage and laterality were more error-prone, particularly in cervix, liver, and lung cases, where these details are often embedded in narrative descriptions or distributed across sections of the report rather than summarized explicitly [34]. Expert reviewers noted that even for human abstractors, accurate extraction of these variables can require inferential reasoning or synthesis of scattered findings. These patterns underscore that model performance is shaped not only by algorithmic capabilities but also by the linguistic and documentation norms of specific organ systems and pathology subspecialties. To be clinically reliable, AI-based extraction frameworks must therefore adapt to these real-world variations in reporting style, especially for use in applications such as cancer registry abstraction and structured synoptic reporting.

Performance on the internal Moffitt dataset, which included brain, breast, and lung pathology reports with both standard variables and organ-specific biomarkers, further demonstrated the robustness of the framework across institutional settings and cancer types. This cohort presented unique challenges due to variability in documentation style, particularly in breast cancer reports, where stage and grade were often embedded in narrative assessments or ancillary sections rather than clearly labeled fields. Biomarker extraction proved particularly challenging, with overall accuracy of 70.6% ± 8.1%, though certain markers like TP53 achieved better accuracy. Despite this complexity, the framework achieved high extraction accuracy for standard variables (88.2% ± 7.2%), underscoring its adaptability to diverse real-world reporting. Statistical analyses revealed highly significant effects across all factors in the TCGA dataset (model: *F* = 1716.82, *p <* 0.001; variable: *F* = 3236.68, *p <* 0.001; organ: *F* = 1946.43, *p <* 0.001), with complex three-way interactions (*F* = 24.74, *p <* 0.001) highlighting the influence of clinical context on model performance. The multi-evaluator approach proved crucial, with evaluator comparison showing significant differences (*F* = 2633.79, *p <* 0.001) despite high correlations (*r >* 0.93), suggesting complementary perspectives in assessment. In the Moffitt dataset, model effects remained strong (*F* = 477.31, *p <* 0.001), while biomarker extraction showed the greatest challenge (*F* = 538.00, *p <* 0.001). The adjudication stage played a critical role in mitigating variability by synthesizing outputs from eleven LLMs and three evaluators, generating consensus predictions based on justification quality and multi-evaluator agreement [35]. This consensus-driven process virtually mirrors the clinical practice of reconciling multiple expert interpretations and supports transparent, auditable decision-making, an essential feature for deploying AI in complex and institution-specific pathology workflows.

Expert review of extracted outputs offered critical insights into the clinical context underlying model performance. Across both the TCGA and Moffitt datasets, pathologists frequently observed that apparent “errors” by the model often stemmed from ambiguity or inconsistency in the source documentation, rather than algorithmic failure. For instance, laterality was flagged as incorrect in cases where it was either not clinically applicable (e.g., cervix, bladder) or where bilateral disease was noted without specifying which side the specimen represented. Stage misclassifications similarly arose from the use of various staging schemes or when reports lacked complete TNM information, such as missing nodal or metastatic status or omitted descriptors like “Mx” or “Nx”, which also challenged expert interpretation. In many of these scenarios, reviewers noted that the model’s output reflected a plausible clinical inference, even if it did not fully align with reporting conventions. These observations underscore that diagnostic accuracy in automated systems must be interpreted in the broader context of real-world variability in documentation practices.

Expert review of the Moffitt dataset (breast, brain, and lung cancers) uncovered organ-specific reasoning challenges that were not evident through automated evaluation alone. In breast cancer reports, pathologists frequently noted that sentinel lymph node status, denoted by markers such as “(sn)” or “(i-)”, was omitted from predicted stage values and inadequately addressed in model justifications. Brain tumor cases revealed difficulty with accurate site and laterality assignment, especially when specimens were labeled differently from surgical targets or when multiple involved sites were present. In lung and metastatic cases, the most common issue was confusion between the biopsy site (e.g., lymph node or liver) and the presumed primary site (e.g., lung), occasionally resulting in under-classification of behavior or grade. Reviewers also observed that even when variable values were correct, model rationales often failed to highlight key clinical cues, limiting transparency and auditability. These findings reinforce the importance of human-in-the-loop validation and suggest that rationale filtering may improve model alignment with expert expectations in complex diagnostic contexts.

A central innovation of this framework lies in its integration of explicit reasoning and multi-evaluator consensus throughout the extraction process. Rather than relying on black-box predictions or a single evaluator, each LLM is prompted to generate not only a structured variable value but also a brief textual justification. These justifications are then evaluated by three independent reasoning models (DeepSeek-R1-large, Qwen3-32B, and QWQ-32B), enabling multiple layers of validation that emphasize coherence and clinical plausibility from diverse perspectives. The high inter-evaluator agreement (correlations > 0.89 across all datasets) confirms the robustness of this approach, while the significant differences between evaluators (p < 0.05) suggest that each contributes unique insights to the assessment process. This approach brings interpretability into a domain where opaque AI outputs are unacceptable and auditability is critical. The final aggregation stage synthesizes responses from eleven models, prioritizing inter-model agreement and justification quality, an approach that mirrors how clinical consensus is reached in pathology through the reconciliation of multiple expert viewpoints. By aligning technical design with established clinical reasoning practices, the system produces outputs that are both reliable and explainable. For users such as pathologists and cancer registrars, this design supports a shift from passive oversight to active validation, where the rationale behind each suggestion is transparent and subject to expert judgment [35].

The performance of extraction models across TCGA and Moffitt datasets is clustered by training recipe rather than parameter count. Models explicitly optimized for step-wise reasoning and strict instruction-following, such as Phi-4-medium, DeepSeek-R1, Llama-3.3, and Tulu-3, consistently outperformed others, reflecting the large-scale curated/synthetic data used in their training and modern post-training techniques (DPO/RL/verification) that improve schema-bound extraction fidelity [36, 37, 38, 39]. By contrast, efficiency-oriented baselines (Mistral-small, Mixtral-medium) emphasize throughput and earlier tuning objectives, which can underperform on format-rigid clinical extraction tasks [40, 41]. Additional contributors to this performance lag among the models include differences in instruction-tuning goals (reasoning/extraction vs general helpfulness) [42], tokenizer/label surface-form compatibility for oncology terms and roman-numeral staging [43, 44], and robustness to domain shift between public (TCGA) and institutional notes. Together, these mechanisms suggest practical selection should be based on the specifically designed criteria for the given reasoning task.

Beyond model performance, the framework is designed with real-world clinical deployment in mind, particularly for environments with stringent data privacy and governance requirements. By utilizing locally hosted, open-source LLMs, the system enables HIPAA-compliant inference within institutional firewalls, avoiding the need to transmit protected health information to external servers [45]. This makes it well-suited for integration into hospital infrastructure, including pathology departments and cancer centers. The structured outputs generated by the adjudication model are readily compatible with downstream tasks such as cancer registry abstraction, synoptic report generation, and clinical trial screening. Each extracted variable is accompanied by a concise rationale, allowing pathology staff to efficiently review and approve outputs in human-in-the-loop workflows. This balance between automation and interpretability is especially critical in oncology, where diagnostic precision has direct implications for treatment, staging, and prognosis. By embedding transparency and alignment with clinical practice into the system design, the framework supports safe, scalable adoption of LLMs in routine pathology workflows.

While this study demonstrates the reliability and clinical relevance of a reasoning-based LLM framework for structured pathology report extraction, several limitations warrant consideration. First, although strong performance was observed across multiple organ sites, the generalizability of the system to rare cancers, atypical documentation formats, or resource-constrained settings remains to be evaluated. Second, the adjudication process, though designed to promote transparency, relies on the reasoning model’s assessment of justification quality, which may introduce its own biases, especially in cases involving ambiguous or incomplete language. Third, expert validation, while essential, was necessarily limited to a subset of reports due to the labor-intensive nature of manual review. Future work will explore the integration of uncertainty estimation, active learning for iterative refinement, expansion to additional variables such as biomarkers, treatment response, or recurrence status, and dedicated pediatric evaluation including curation of pediatric synoptic templates in collaboration with pediatric pathologists. Prospective validation within live clinical environments, including deployment into laboratory information systems and registry pipelines, will be critical for assessing operational impact on accuracy, workflow efficiency, and downstream decision-making [29]. Addressing these challenges will be key to realizing the full potential of this framework as a scalable, trustworthy foundation for AI-enabled pathology practice.

## Data Availability

Some data produced in the present study are available upon reasonable request to the authors.

## Author Contributions

AT and AW contributed equally as co-first authors. AT and AW designed and implemented the three-stage extraction framework, conducted all computational analyses, and performed statistical evaluations. EU, AK, FK, WSC, ZGO, DSV, and MMB served as expert evaluators, independently reviewing and scoring model outputs from pathology reports. KV assisted with manuscript preparation and revision. MBS provided expertise in cancer epidemiology and study design. GR supervised the project as principal investigator, provided guidance on framework development, and secured funding. AT, AW, and GR drafted the manuscript. All authors reviewed, edited, and approved the final manuscript.

## Data Availability Statement

The Cancer Genome Atlas (TCGA) pathology reports used in this study are publicly available through the National Cancer Institute’s Genomic Data Commons (GDC) portal. The Moffitt Cancer Center dataset contains de-identified patient information and is not publicly available due to institutional data governance policies and patient privacy protections. Access to the Moffitt dataset may be requested through the corresponding author and is subject to institutional review board approval and execution of a data use agreement. Code for the extraction framework and analysis pipelines will be made available upon reasonable request to the corresponding author.

## Ethics Approval and Consent to Participate

This study utilized two datasets: (1) publicly available pathology reports from The Cancer Genome Atlas (TCGA), and (2) a de-identified internal dataset from Moffitt Cancer Center. The TCGA dataset is publicly accessible and does not contain individually identifiable information; therefore, informed consent was not required. Use of the internal Moffitt dataset was approved under a protocol with a waiver of informed consent granted by the Institutional Review Board (IRB) at Moffitt Cancer Center, in accordance with institutional policies and federal regulations for research involving de-identified data.

## Appendix A

**Sample JSON, Raw Report, and LLM Reasoning Output**

## Appendix B

**System Prompts**

